# Association of Rare *APOE* Missense Variants R189C and W276C With Risk of Alzheimer Disease

**DOI:** 10.64898/2026.05.08.26352778

**Authors:** Yann Le Guen, Dylan Reil, Rosemary J. Jackson, Andrés Peña-Tauber, Chaitan Khosla, Michael D. Greicius

## Abstract

Rare *APOE* missense variants in admixed populations can clarify mechanisms of Alzheimer disease (AD) risk beyond ε2 and ε4. We analyzed *APOE* coding variation in the Alzheimer disease Sequencing Project and AllofUs and meta-analyzed ancestry-adjusted case-control associations. Two ε3-linked variants enriched in Native American ancestry among Admixed American individuals were associated with AD: R189C increased risk (odds ratio (OR), 3.35; 95% confidence interval (CI), 1.25–9.00; *P* = 0.016), whereas W276C was protective (OR, 0.28; 95% CI, 0.10–0.82; *P* = 0.020). In a nascent ApoE secretion assay that resolves high molecular weight lipid-bound species from lipid-poor species, R189C showed an *APOE-*ε4-like shift toward the lipid-bound fraction. In an orthogonal self-association assay, W276C reduced ApoE self-association to the protective *APOE*-Jacksonville variant level. These findings expand the spectrum of *APOE* missense variants that are associated with AD and implicate C-terminal ApoE conformation and lipidation states as tractable mechanisms for pathogenesis and therapeutic targeting.

## Main text

*APOE* remains the strongest common genetic determinant of late-onset Alzheimer disease (AD)^1^, but the molecular basis of its effect is still unresolved. Beyond the canonical ε2 and ε4 alleles, rare coding variants offer a powerful way to connect human genetics to specific APOE protein states. This has become especially important as *APOE* is increasingly viewed not only as a risk locus, but also as a mechanistically actionable therapeutic axis in AD^2^. Consistent with this view, recent human genetic evidence indicates that *APOE* loss-of-function variants are compatible with longevity and may be associated with resistance to AD-related pathology^3^, supporting the idea that ε4 risk is driven at least in part by gain of abnormal function rather than by a reduction in normal APOE activity.

Recent human genetic studies have begun to widen the array of *APOE* missense variants associated with AD. In African ancestry populations, the ε3-linked R145C variant increases AD risk in a genotype-specific manner^4^. In predominantly European datasets, the C-terminal variants V236E (Jacksonville) on ε3 and R251G on ε4 are associated with substantially lower AD risk^5^, supporting an important role for the carboxyl terminus of apoE in disease biology. The Christchurch variant R136S drew broad attention after a homozygous carrier in a PSEN1 E280A kindred showed a striking delay in age-at-onset of symptoms^6^, and a subsequent report suggested delayed onset even in heterozygotes in that kindred^7^. The heterozygote finding has been contended^8^, however, underscoring the value of population-based statistical genetics over exceptional-case reports.

Here we asked whether additional *APOE* missense variants enriched in Admixed American populations can be added to the array of AD-associated missense variants. We analyzed seven non-synonymous *APOE* variants selected on the basis of representation in the Admixed American group in gnomAD and observed in the Admixed American subsets of two large sequencing resources: the Alzheimer’s Disease Sequencing Project (ADSP) and All of Us whole-genome sequencing. In ADSP, ancestry was assigned with SNPWeights^9^ using 1000 Genomes reference populations^10^; in All of Us, we used the published genetic-ancestry framework based on projection into a reference principal-component space and classifier-based assignment^11^. Association analyses were performed within the Admixed American subset and combined by inverse-variance weighted meta-analysis (**Fig. 1 and Extended Data Fig. 1, and Extended Data Table 1**).

**Fig. 1.**
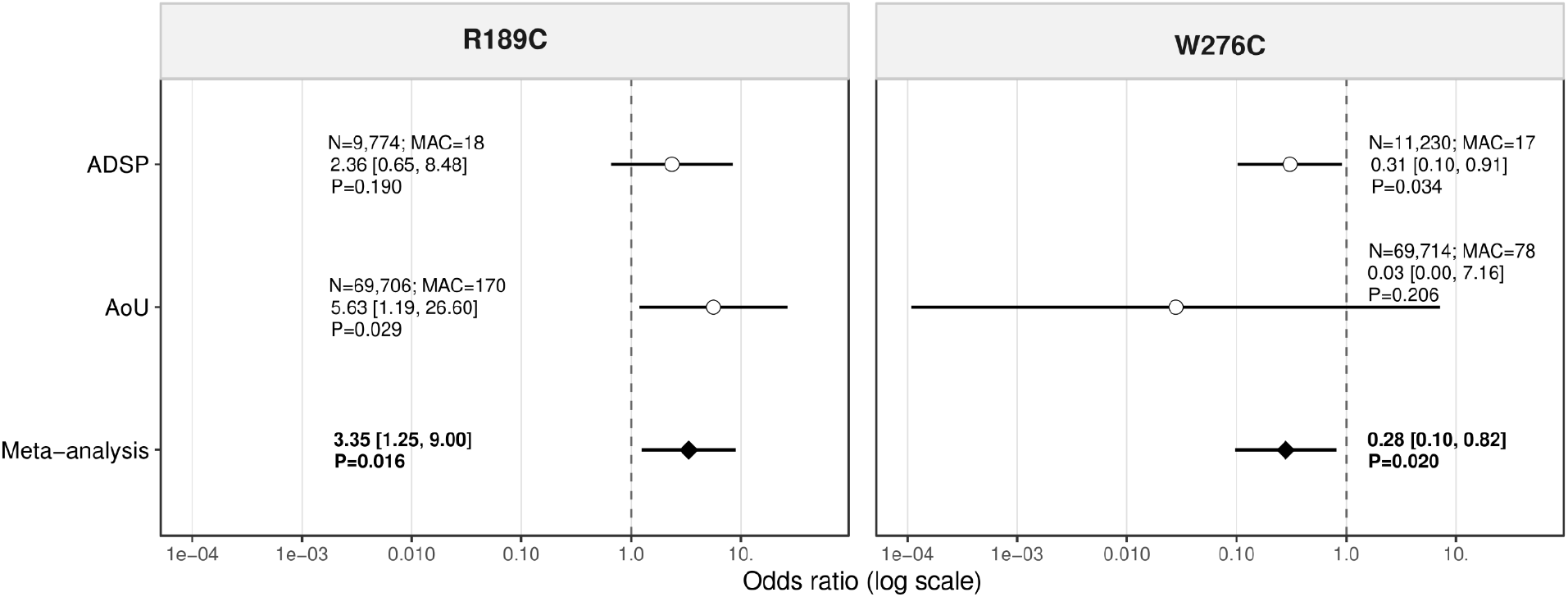
Genetic association of *APOE* R189C and W276C with Alzheimer disease in Admixed American participants. Forest plots showing cohort-specific and fixed-effect meta-analysis estimates for R189C and W276C in ADSP and All of Us. Circles indicate cohort-specific odds ratios, horizontal lines indicate 95% confidence intervals and diamonds indicate pooled fixed-effect estimates. The vertical dashed line marks an odds ratio of 1. Sample size and minor allele count (MAC) for each cohort are shown in the figure. All of Us used a proxy-AD phenotype.

Two ε3-linked C-terminal variants showed evidence of association with AD across both datasets. R189C was associated with increased risk in the combined analysis (odds ratio (OR) = 3.35, 95% confidence interval (CI) = 1.25–9.00, P = 0.016), with a directionally consistent signal in both ADSP (OR = 2.36) and All of Us (OR = 5.63). W276C showed the opposite pattern and was associated with reduced risk (OR = 0.28, 95% CI = 0.10–0.82, P = 0.020), again with concordant protective point estimates in ADSP (OR = 0.31) and All of Us (OR = 0.03). These effects were detected despite very low allele counts, emphasizing both the rarity of the variants and the value of aggregating diverse cohorts. The remaining tested variants did not reach nominal significance in the combined analysis.

One point deserves emphasis. In this Admixed American meta-analysis, R136S (Christchurch) showed no evidence of protection and instead had a non-significant point estimate above the null (OR = 1.42, 95% CI 0.18–11.25, P = 0.740). This does not directly contradict observations in autosomal-dominant AD kindreds^6,7^, where genetic background, ascertainment and mechanism may differ, but it argues against assuming that Christchurch is uniformly protective across clinical and ancestral contexts. More broadly, it serves as a reminder that case reports, even when supported by mechanistic studies, are not sound proxies for population-based statistical associations.

To connect these associations to ApoE biology, we evaluated R189C and W276C in two orthogonal functional assays (**Fig. 2**). The first was an ApoE secretion assay developed in mammalian cells, in which secreted ApoE resolves into a high molecular weight, lipid-bound lipoprotein particle (previously termed Peak 1) and a lower molecular weight, lipid-poor species (previously termed Peak 2)^12^. The former particle not only elicits cholesterol efflux at a higher rate from cells with ATP-dependent lipid pump activity but it also undergoes more effective receptor mediated endocytosis. Relative to ApoE3, expression of both ApoE4 and ApoE3-R189C led to more of the high molecular weight ApoE species (Tukey-adjusted P = 0.0012 for each comparison). In contrast, ApoE3-R189C was not detectably different from ApoE4 (P = 0.9995) and ApoE4-R251G yielded less of the high molecular weight species than ApoE4 (P = 0.0031). These results suggest that R189C mimics ApoE4 in its lipid secretion characteristics.

**Fig. 2.**
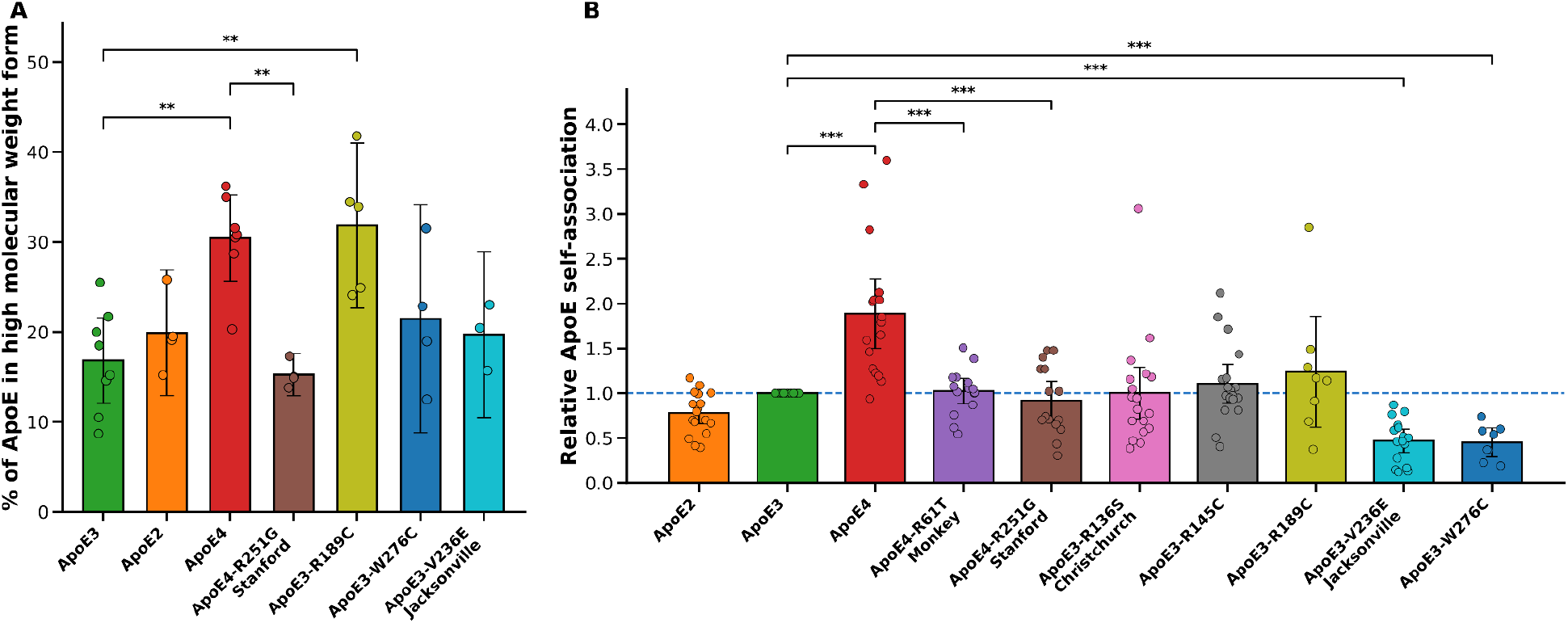
Orthogonal functional assays implicate distinct ApoE states for R189C and W276C. **A)** Percentage of total secreted ApoE recovered in the high molecular weight, lipid-bound form in the ApoE secretion assay. Global differences were assessed by one-way ANOVA and selected pairwise comparisons were evaluated with Tukey’s multiple-comparisons test. **B)** Relative maximum luminescence intensity in the split-luciferase ApoE self-association assay under serum-free conditions. W276C shows reduced self-association, resembling the protective Jacksonville variant V236E. Global differences were assessed by Kruskal–Wallis testing, and selected pairwise comparisons were evaluated using two-sided Mann–Whitney U tests with Holm correction. Points denote individual measurements; bars and error bars indicate mean ± 95% confidence interval. *P < 0.05, **P < 0.01, ***P < 0.001.

The second assay, developed independently, measured ApoE self-association using a cell-based association readout^13^. In prior work^14^, the Jacksonville variant V236E was shown to reduce ApoE self-aggregation, providing a plausible biochemical basis for protection. In our assay, ApoE4 showed higher self-association than ApoE3 (Holm-adjusted P = 2.05 × 10^-5^). Both ApoE3-V236E and ApoE3-W276C showed less self-association than ApoE3 (Holm-adjusted P = 1.35 × 10^-6^ and P = 1.65 × 10^-5^, respectively), and ApoE4-R251G also showed less self-association than ApoE4 (Holm-adjusted P = 7.92 × 10^-4^). ApoE3-V236E and ApoE3-W276C were not detectably different from one another (two-sided Mann–Whitney P = 0.93). In contrast, ApoE3-R189C did not show this Jacksonville-like profile. In contrast, ApoE3-R189C did not show this Jacksonville-like profile. Thus, the two newly implicated Admixed American variants have distinct intermolecular interaction characteristics than ApoE3: R189C has an ApoE4-like preference for lipidation, and W276C has a Jacksonville-like reduction in self-association.

These findings extend the spectrum of AD-associated *APOE* missense variants in a population that has been underrepresented in AD genetics and point again to the C-terminal region of APOE as a key determinant of disease risk. More broadly, they reinforce the idea that substitutions that introduce or remove Cys residues are unusually consequential in *APOE* pathobiology: the two common alleles with the strongest effects on AD risk are themselves defined by Cys/Arg changes, with *APOE*-ε2 introducing a cysteine residue at position 158 and *APOE*-ε4 lacking the Cys present at residue 112 in *APOE*-ε3. Against this background, it is notable that both R189C and W276C introduce cysteine residues onto an ε3 background, yet they associate with opposite clinical effects. This argues against a simple model in which Cys addition uniformly confers protection or risk, and instead suggests that residue position and its impact on intermolecular interactions with lipids and/or proteins are critical. That interpretation is consistent with prior human genetics ^5^, with biochemical data mapping lipid binding and oligomerization properties of ApoE to its C-terminal domain and with other data linking the physical state of ApoE to its role in amyloid handling, tau-mediated neurodegeneration^15^ and receptor-dependent trafficking^16^.

Our study has limitations. The variants described here are very rare, confidence intervals remain wide, and the All of Us analysis relied in part on a proxy phenotype. The functional experiments were designed to test directionally informative biochemical states rather than establish a full causal chain from variant to disease. Even so, the concordance between human genetics and orthogonal assays strengthens the central conclusion: rare *APOE* coding variants in Admixed American populations can reveal mechanistically distinct routes to AD risk and resilience. Together, these data support a model in which C-terminal ApoE biology, including lipidation and self-association, is not merely correlative but is closely tied to pathogenesis and may inform future *APOE*-directed therapeutic strategies^2,17^.

## Online Methods

### Study design and cohorts

We performed a two-cohort genetic association study of non-synonymous *APOE* variants in Admixed American participants from ADSP and All of Us. In both cohorts, the Admixed American designation corresponds to populations from the Americas with mixed Native American, European and African ancestry, as represented in the 1000 Genomes Project by groups including MXL (Mexican Ancestry from Los Angeles, USA), PUR (Puerto Ricans from Puerto Rico), CLM (Colombians from Medellín, Colombia) and PEL (Peruvians from Lima, Peru)^10^. In ADSP, ancestry was assigned with SNPWeights,^9^ and the Admixed American subgroup for analysis was defined as non-duplicated case-control participants from merged whole-genome and whole-exome sequencing data with estimated AMR ancestry proportion >15%. ADSP cases had clinically adjudicated Alzheimer disease and controls were cognitively unimpaired (**Extended Data Table 2**). In All of Us, Admixed American assignment followed the published genetic-ancestry framework^11^, based on projection into reference principal-component space and classifier-based assignment using the same 1000 Genomes reference groups (**Extended Data Table 3**). Cases were defined by the presence of an electronic health record diagnosis of Alzheimer disease (ICD-10 code G30). Proxy cases were defined by a self-reported first-degree family history of dementia/Alzheimer disease in a mother, father or sibling^18,19^. Controls were defined by the absence of both criteria.

### Variant selection

We selected *APOE* missense variants *a priori* from the Admixed American group on the basis of representation in gnomAD and carrier counts sufficient for association testing in the available sequencing datasets. The seven analyzed variants were signal peptide/T11A, E13K, R136S, R189C, V195G, R260C and W276C. Variant nomenclature in Extended Data is shown in both mature-protein and precursor numbering where relevant.

### Association testing in ADSP

Within ADSP Admixed American participants, we tested association between each *APOE* variant and case-control status using a generalized linear mixed-effects model with a binomial outcome, following the framework used in prior ADSP *APOE* rare-variant analyses^4,5^, Models were adjusted for sex, *APOE*-ε2 dosage, *APOE*-ε4 dosage and 3 ancestry principal components. Relatedness was accounted for using a sparse genetic relationship matrix. All tests were two-sided. The analyses were implemented in R (v4.5) with the GENESIS package^20^ (v2.40.0) implementing PCRelate/PCAir. The principal components were computed with PCAir^21^ on the subset of Admixed Americans participants merged from ADSP whole-exome (WES) and whole-genome sequencing (WGS) (**Extended Data Figure 2**).

### Association testing in All of Us

Within All of Us Admixed American participants, we tested association between each APOE variant and proxy Alzheimer/Dementia disease status. Cases and proxy-cases were defined by the presence of either an electronic health record diagnosis of Alzheimer disease (ICD-10 code G30) or a self-reported first-degree family history of dementia/Alzheimer disease in a mother, father or sibling; controls were defined by the absence of both criteria. We restricted analyses to an unrelated, QC-passing set by excluding samples flagged in the All of Us ancillary files relatedness_flagged_samples.tsv and flagged_samples.tsv. Association testing was performed with PLINK v2.0.0-a.6.11LM using --glm firth-residualize, adjusting for age, sex, *APOE*-ε2 dosage, *APOE*-ε4 dosage and 16 genetic principal components precomputed by All of Us^11^.

### Meta-analysis

We combined ADSP and All of Us estimates using inverse-variance weighted fixed-effect meta-analysis on the log-odds-ratio scale. For the proxy-enriched All of Us analysis, effect sizes and standard errors were doubled before meta-analysis to account for the use of proxy phenotypes. P values were two-sided.

### ApoE secretion assay

Functional studies were performed in a mammalian-cell expression system in which secreted, recombinant ApoE was purified and resolved by size-exclusion chromatography into a high molecular weight, lipid-bound fraction (previously termed Peak 1) and a lower molecular weight, lipid-poor fraction (previously termed Peak 2). Prior work showed that the former fraction has greater cholesterol efflux activity in assays targeted at testing the ability of ApoE to stimulate ATP-dependent lipid export; it also undergoes more effective receptor-mediated endocytosis than the lipid-poor species^12^. The primary readout in the present study was the percentage of the total secreted ApoE recovered in the high molecular weight fraction.

Expi293F cells were maintained in Expi293 Expression Medium (Thermo Fisher Scientific, A1435101) at 37 °C and 8% CO2 with shaking at 120 r.p.m. ApoE mutant constructs in a pCMV4 backbone were transfected at 3 × 10^6^ cells/ml using ExpiFectamine 293 (Thermo Fisher Scientific, A14525). After 4 d, the conditioned medium was cleared by centrifugation, filtered and applied to a HiTrap Heparin HP column (Cytiva, 17040701) on an ÄKTA Pure system. ApoE-containing fractions were identified by SDS–PAGE, pooled, concentrated and further separated by size-exclusion chromatography on a HiLoad 16/600 Superdex 200 pg column (Cytiva, 28989335). The two distinct ApoE fractions were pooled separately. ApoE concentration in the low molecular weight fraction was measured by absorbance at 280 nm; ApoE concentration in the high molecular weight fraction was quantified by immunoblotting against a low molecular weight ApoE standard using anti-ApoE antibody 16H22L18 (Thermo Fisher Scientific, 701241). Constructs encoding ApoE2, ApoE3, ApoE4, ApoE4-R251G, ApoE3-R189C, ApoE3-W276C and ApoE3-V236E were analyzed under matched conditions.

For statistical analysis, distributional assumptions were evaluated by Shapiro–Wilk testing within groups and homogeneity of variance was assessed using Levene’s test centered on the median. Global differences across constructs were assessed using two-sided one-way ANOVA. When the omnibus test was significant, pairwise differences were evaluated using Tukey’s multiple-comparisons test. The comparisons emphasized in Fig. 2a were prespecified on biological grounds and included ApoE3 versus ApoE4, ApoE4 versus ApoE4-R251G, ApoE3 versus ApoE3-W276C, and ApoE3 versus ApoE3-R189C. Individual points represent measurements from individual experiments, and bars with error bars indicate mean ± 95% confidence interval. A two-sided P value < 0.05 was considered statistically significant.

### Split-luciferase self-association assay

ApoE self-association was assessed in a split-luciferase assay, as previously described^13^. In brief, plasmids were engineered to express ApoE variants fused at the C terminus to either the Small BiT (SmBiT) or Large BiT (LgBiT) luciferase fragment. HEK293 cells were transiently transfected with Lipofectamine 2000 and the corresponding ApoE-SmBiT or ApoE-LgBiT constructs. After 4–6 h, the medium was replaced with Gibco Opti-MEM Reduced Serum Medium supplemented with 1% (v/v) penicillin–streptomycin and 1% (v/v) GlutaMAX, and cells were incubated for 48 h under standard culture conditions. This 48-h serum-free condition was used because longer incubations increased apoE cleavage and serum reduced isoform-specific differences.

Conditioned medium was collected and centrifuged at 10,000 r.p.m. for 3 min to remove cellular debris. ApoE abundance in the medium was assessed by immunoblotting to enable concentration normalization across variants. Membranes were incubated overnight at 4 °C with primary antibody against ApoE (Novus Biologicals, NBP1-31123; 1:2,000 in milk), washed, and then incubated with IRDye 800CW donkey anti-rabbit secondary antibody (LI-COR, 926-32213; 1:5,000 in 1× TBS) for approximately 1 h. Membranes were imaged on an Odyssey CLx system, and band intensities were quantified using Image Studio 6.0 (LI-COR). ApoE concentrations were then normalized to the sample with the lowest signal.

ApoE abundance in conditioned medium was assessed by immunoblotting, and samples were normalized to equal relative ApoE input across constructs on the basis of band intensity before assay assembly. Following this normalization, 20 μl of SmBiT-tagged apoE and 20 μl of LgBiT-tagged ApoE were combined for each homodimer measurement. Absolute ApoE concentrations were not determined; accordingly, the assay provides a relative self-association readout across matched conditions. Luciferase substrate was prepared as 0.4 μl HiBiT Lytic Substrate in 59.6 μl of 0.25% Pluronic acid and 20 μM HEPES in HBSS (+/+), and added per well. Luminescence was measured after 45 min using a GloMax Navigator microplate luminometer (Promega). Relative maximum luminescence intensity was used as the self-association readout and was normalized to ApoE3 within each experimental run. Constructs included ApoE2, ApoE3, ApoE4, ApoE4-R61T, ApoE4-R251G, ApoE3-R136S, ApoE3-R145C, ApoE3-R189C, ApoE3-V236E and ApoE3-W276C.

For the split-luciferase assay, outlier exclusion was prespecified at the construct level using a robust modified z-score based on the median absolute deviation, with |modified z| > 3.5 considered outlying. One ApoE4-R251G measurement and one ApoE3-R145C measurement met this criterion. The ApoE4-R251G point also corresponded to a documented technical artifact during the assay. Analyses were robust to inclusion of all data points, and sensitivity analyses with and without the excluded points did not alter the interpretation of the prespecified biological comparisons.

For statistical analysis, global differences across constructs were assessed using a two-sided Kruskal–Wallis test. Because the ApoE3 reference condition was normalized to 1.0, yielding minimal within-group variance in that reference group, pairwise inference for the figure was based on prespecified two-sided Mann–Whitney U tests with Holm correction for multiple comparisons. The planned comparisons included ApoE3 versus ApoE2, ApoE3 versus ApoE4, ApoE4 versus ApoE4-R61T, ApoE4 versus ApoE4-R251G, ApoE3 versus ApoE3-R136S, ApoE3 versus ApoE3-R145C, ApoE3 versus ApoE3-R189C, ApoE3 versus ApoE3-V236E and ApoE3 versus ApoE3-W276C. The comparisons annotated in Fig. 2b were the biologically prioritized subset of these planned tests. Individual points represent measurements from individual experiments, and bars with error bars indicate mean ± 95% confidence interval. A two-sided adjusted P value < 0.05 was considered statistically significant.

## Data Availability

ADSP data are available through NIAGADS. AllOfUs data are available through the AllOfUs workbench.

https://dss.niagads.org/

https://workbench.researchallofus.org/

---

**Extended Data Figure 1.**
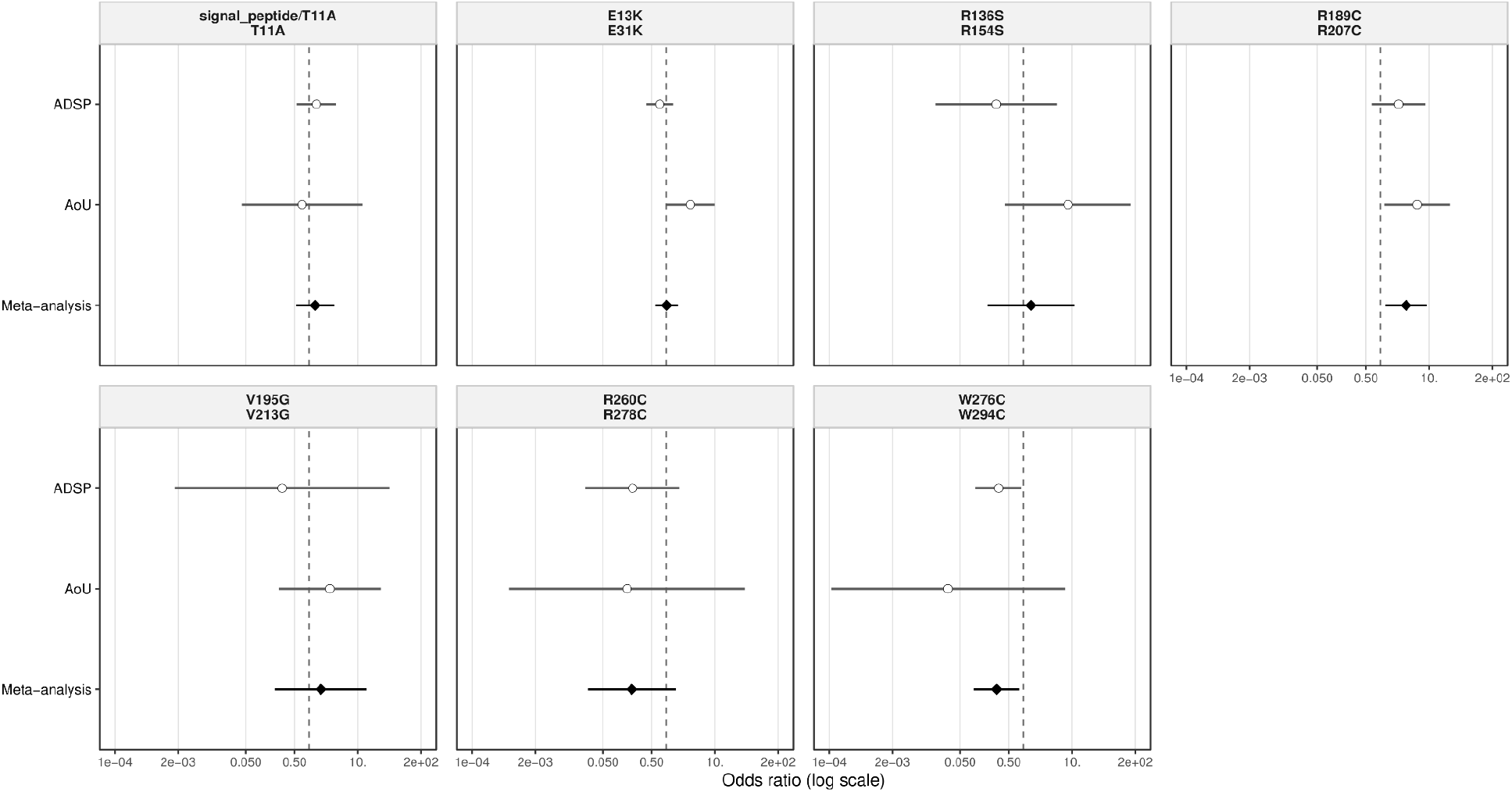
Meta-analysis of seven *APOE* missense variants tested in the Admixed American subset. Forest plots showing ADSP, All of Us and fixed-effect meta-analysis estimates for the seven analyzed *APOE* variants: signal peptide/T11A, E13K, R136S, R189C, V195G, R260C and W276C.

**Extended Data Table 1.** Cohort-specific and meta-analysis association results for seven *APOE* missense variants in Admixed American participants.

| Variant | ADSP |  |  |  | AoU |  |  |  | Meta-analysis |  |  |  |
| --- | --- | --- | --- | --- | --- | --- | --- | --- | --- | --- | --- | --- |
|  | N | MAC | OR [95% CI] | p-value | N | MAC | OR [95% CI] | p-value | N | MAC | OR [95% CI] | p-value |
| signal_peptide/T11A (T11A) | 12,018 | 23 | 1.41 [0.55, 3.63] | 0.4734 | 69,699 | 56 | 0.72 [0.04, 12.41] | 0.8179 | 81,717 | 79 | 1.32 [0.54, 3.23] | 0.5429 |
| E13K (E31K) | 11,991 | 51 | 0.73 [0.39, 1.37] | 0.3334 | 69,713 | 223 | 3.14 [0.99, 9.93] | 0.0518 | 81,704 | 274 | 1.02 [0.59, 1.77] | 0.9392 |
| R136S (R154S) | 12,016 | 3 | 0.28 [0.02, 4.88] | 0.3792 | 69,708 | 22 | 8.25 [0.42, 162.41] | 0.1650 | 81,724 | 25 | 1.42 [0.18, 11.25] | 0.7400 |
| R189C (R207C) | 9,774 | 18 | 2.36 [0.65, 8.48] | 0.1899 | 69,706 | 170 | 5.63 [1.19, 26.60] | 0.0292 | 79,480 | 188 | 3.35 [1.25, 9.00] | 0.0165 |
| V195G (V213G) | 9,784 | 1 | 0.28 [0.00, 45.11] | 0.6212 | 69,608 | 55 | 2.65 [0.24, 29.49] | 0.4282 | 79,392 | 56 | 1.75 [0.20, 15.48] | 0.6137 |
| R260C (R278C) | 9,784 | 4 | 0.20 [0.02, 1.88] | 0.1601 | 69,715 | 43 | 0.16 [0.00, 41.72] | 0.5150 | 79,499 | 47 | 0.20 [0.02, 1.55] | 0.1221 |
| W276C (W294C) | 11,230 | 17 | 0.31 [0.10, 0.91] | 0.0337 | 69,714 | 78 | 0.03 [0.00, 7.16] | 0.2061 | 80,944 | 95 | 0.28 [0.10, 0.82] | 0.0199 |

**Extended Data Table 2.**
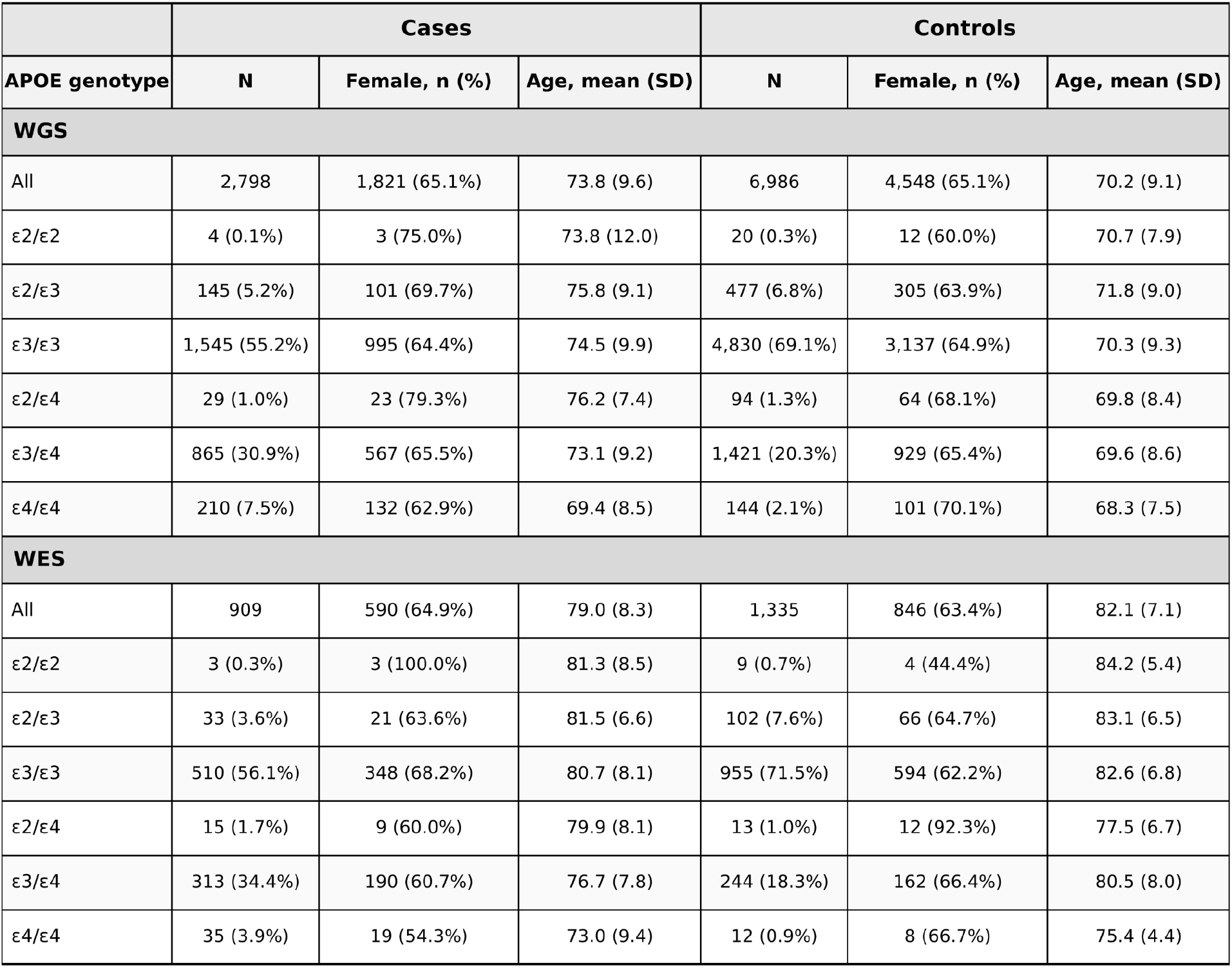
Demographic characteristics of Admixed American Alzheimer’s disease cases and controls in the Alzheimer’s Disease Sequencing Project, stratified by sequencing subset and *APOE* genotype. Demographic summary of ADSP participants included in the Admixed American analysis, shown separately for whole-genome sequencing (WGS) and whole-exome sequencing (WES) subsets and stratified by *APOE* genotype. For each stratum, the table reports sample size, female sex, and age for Alzheimer’s disease cases and controls. Percentages in genotype-specific rows are relative to the corresponding WGS or WES case/control total. Age is reported as mean (s.d.).

**Extended Data Table 3.** Demographic characteristics of Admixed American case/proxy-case and control participants in the All of Us Research Program, stratified by APOE genotype. Demographic summary of All of Us participants included in the Admixed American analysis, stratified by APOE genotype. For each stratum, the table reports sample size, female sex, and age for case/proxy-case and control participants. Percentages in genotype-specific rows are relative to the corresponding All of Us case/proxy-case or control total. Age is reported as mean (s.d.).

|  |  | Cases |  |  | Controls |  |
| --- | --- | --- | --- | --- | --- | --- |
| APOE genotype | N | Female, n (%) | Age, mean (SD) | N | Female, n (%) | Age, mean (SD) |
| All | 2,006 | 1,401 (69.8%) | 60.1 (12.8) | 67,709 | 43,971 (64.9%) | 47.5 (15.7) |
| ε2/ε2 | 3 (0.1%) | 3 (100.0%) | 57.6 (22.1) | 137 (0.2%) | 95 (69.3%) | 49.5 (16.2) |
| ε2/ε3 | 131 (6.5%) | 96 (73.3%) | 59.2 (13.4) | 4,722 (7.0%) | 3,075 (65.1%) | 47.9 (16.0) |
| ε3/ε3 | 1,324 (66.0%) | 937 (70.8%) | 60.5 (12.8) | 47,626 (70.3%) | 30,895 (64.9%) | 47.6 (15.7) |
| ε2/ε4 | 26 (1.3%) | 19 (73.1%) | 59.2 (13.8) | 690 (1.0%) | 463 (67.1%) | 47.9 (15.6) |
| ε3/ε4 | 492 (24.5%) | 324 (65.9%) | 59.5 (12.9) | 13,505 (19.9%) | 8,761 (64.9%) | 47.0 (15.5) |
| ε4/ε4 | 30 (1.5%) | 22 (73.3%) | 60.2 (8.6) | 1,029 (1.5%) | 682 (66.3%) | 46.6 (15.2) |

**Extended Data Figure 2.**
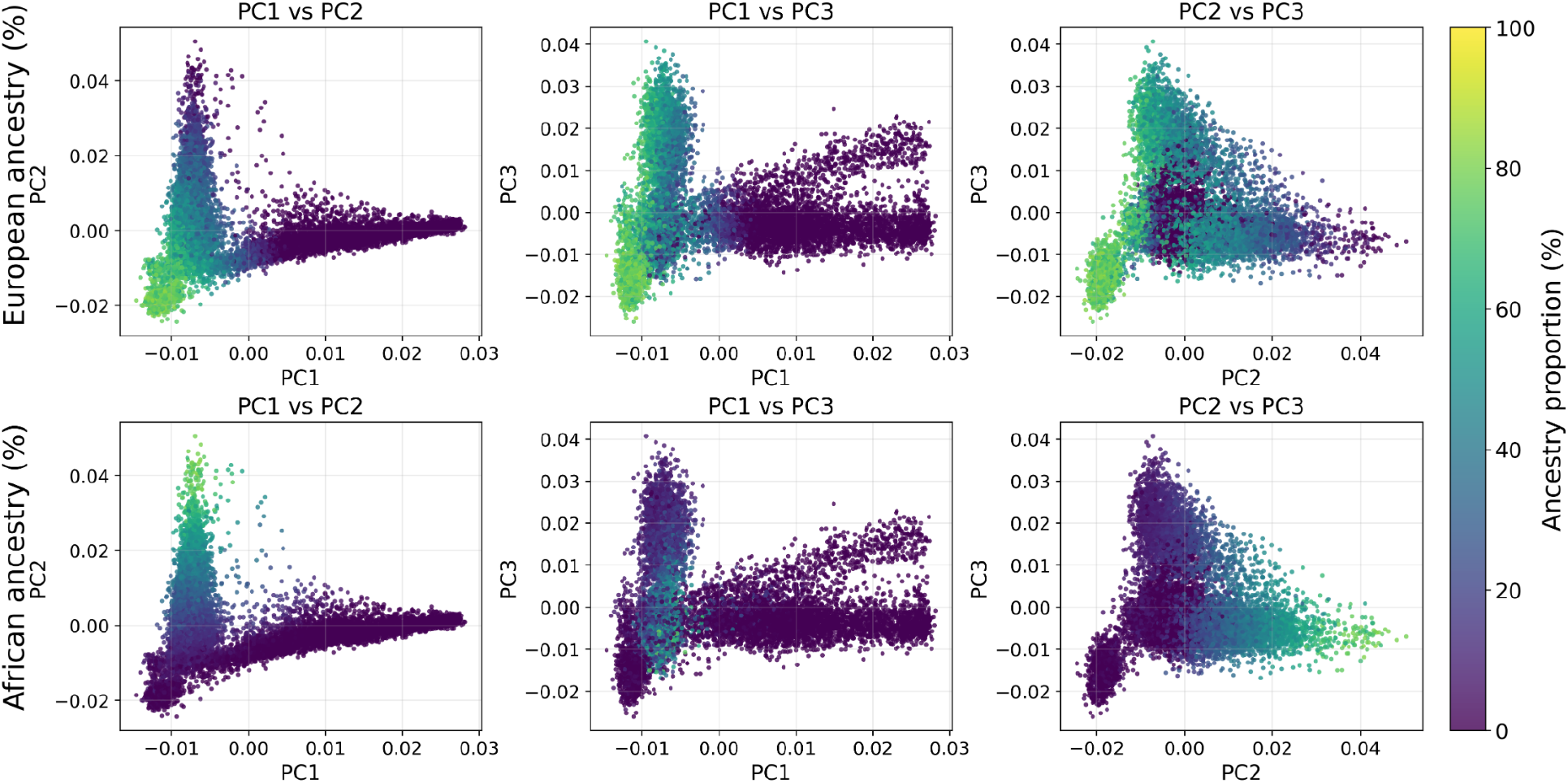
Population structure of the ADSP WGS+WES Admixed American subset. Pairwise plots of the first three principal components (PC1–PC3) are shown for non-duplicated ADSP case-control participants from merged WGS and WES data with estimated AMR ancestry proportion >15%. Principal components were derived using the GENESIS/PCAir pipeline. Samples are colored by estimated EUR ancestry proportion (top row) or AFR ancestry proportion (bottom row), as inferred with SNPweights using projection onto 1000 Genomes reference weights.

## References

1. Belloy, M. E. et al. APOE Genotype and Alzheimer Disease Risk Across Age, Sex, and Population Ancestry. JAMA Neurol. 80, 1284–1294 (2023).

2. Raulin, A.-C. et al. ApoE in Alzheimer’s disease: pathophysiology and therapeutic strategies. Mol. Neurodegener. 17, 72 (2022).

3. Chemparathy, A. et al. *APOE* loss-of-function variants: Compatible with longevity and associated with resistance to Alzheimer’s disease pathology. Neuron 112, 1110–1116.e5 (2024).

4. Le Guen, Y. et al. Association of African Ancestry–Specific APOE Missense Variant R145C With Risk of Alzheimer Disease. JAMA 329, 551–560 (2023).

5. Le Guen, Y. et al. Association of Rare APOE Missense Variants V236E and R251G With Risk of Alzheimer Disease. JAMA Neurol. 79, 652–663 (2022).

6. Arboleda-Velasquez, J. F. et al. Resistance to autosomal dominant Alzheimer’s disease in an APOE3 Christchurch homozygote: a case report. Nat. Med. 25, 1680–1683 (2019).

7. Quiroz, Y. T. et al. APOE3 Christchurch Heterozygosity and Autosomal Dominant Alzheimer’s Disease. N. Engl. J. Med. 390, 2156–2164 (2024).

8. Cochran, J. N., Greicius, M. D. & Goate, A. M. APOE3 Christchurch Heterozygosity and Autosomal Dominant Alzheimer’s Disease. N. Engl. J. Med. 391, 1660–1661 (2024).

9. Chen, C. Y. et al. Improved ancestry inference using weights from external reference panels. Bioinformatics 29, 1399–1406 (2013).

10. Auton, A. et al. A global reference for human genetic variation. Nature 526, 68–74 (2015).

11. Bick, A. G. et al. Genomic data in the All of Us Research Program. Nature 627, 340–346 (2024).

12. Hernandez Arriaza, R. et al. ApoE is Secreted as a Lipid Nanoparticle by Mammalian Cells: Implications for Alzheimer’s Disease Pathogenesis. Biochemistry 64, 4387–4399 (2025).

13. Andrieieva, D. et al. APOE Isoform-Dependent Self-Association Measured by a Split-Luciferase Complementation Assay: Differential Effects of Disease-Risk and Protective Variants. medRxiv 2026.05.09.26352797 (2026) doi:10.64898/2026.05.09.26352797.

14. Liu, C.-C. et al. APOE3-Jacksonville (V236E) variant reduces self-aggregation and risk of dementia. Sci. Transl. Med. 13, eabc9375 (2021).

15. Shi, Y. et al. ApoE4 markedly exacerbates tau-mediated neurodegeneration in a mouse model of tauopathy. Nature 549, 523–527 (2017).

16. Guo, J. L. et al. Decreased lipidated ApoE-receptor interactions confer protection against pathogenicity of ApoE and its lipid cargoes in lysosomes. Cell 188, 187–206.e26 (2025).

17. Vance, J. M. et al. Report of the APOE4 National Institute on Aging/Alzheimer Disease Sequencing Project Consortium Working Group: Reducing APOE4 in Carriers is a Therapeutic Goal for Alzheimer’s Disease. Ann. Neurol. 95, 625–634 (2024).

18. Liu, J. Z., Erlich, Y. & Pickrell, J. K. Case-control association mapping by proxy using family history of disease. Nat. Genet. 49, 325–331 (2017).

19. Le Guen, Y. et al. Population-scale burden analysis of rare damaging coding variants identifies novel risk genes for Alzheimer’s disease and Parkinson’s disease. 2026.03.03.26347540 Preprint at 10.64898/2026.03.03.26347540 (2026).

20. Gogarten, S. M. et al. Genetic association testing using the GENESIS R/Bioconductor package. Bioinformatics 35, 5346–5348 (2019).

21. Conomos, M. P., Miller, M. B. & Thornton, T. A. Robust Inference of Population Structure for Ancestry Prediction and Correction of Stratification in the Presence of Relatedness. Genet. Epidemiol. 39, 276–293 (2015).

